# Close, But no Cigar: Comparative Evaluation of ChatGPT-4o and OpenAI o1-preview in Answering Pancreatic Ductal Adenocarcinoma-Related Questions

**DOI:** 10.1101/2025.07.26.25332239

**Authors:** Cheng-Peng Li, Yuan Chu, Dao-Ning Liu, Erfan Ghanad, Schaima Abdelhadi, Flavius Șandra-Petrescu, Christoph Reißfelder, Georgi Vassilev, Cui Yang

## Abstract

**Background:** This study aimed to evaluate the effectiveness of ChatGPT-4o and OpenAI o1-preview in responding to pancreatic ductal adenocarcinoma (PDAC)-related queries. The study assessed both LLMs’ accuracy, comprehensiveness, and safety when answering clinical questions, based on the National Comprehensive Cancer Network® (NCCN) Clinical Practice Guidelines for PDAC.

**Methods:** The study used a 20-question dataset derived from clinical scenarios related to PDAC. Two board-certified surgeons independently evaluated the responses by ChatGPT-4o and OpenAI o1-preview for their accuracy, comprehensiveness, and safety using a Likert scale. Statistical analyses were conducted to compare the performances of the two models. We also analyzed the impact of OpenAI o1-preview’s Chain of Thought (CoT) technology.

**Results:** Both models demonstrated high median scores across all dimensions (5 out of 5). OpenAI o1-preview outperformed ChatGPT-4o in comprehensiveness (p = 0.026) and demonstrated superior reasoning ability, with a higher accuracy rate of 75% compared to 60% for ChatGPT-4o. OpenAI o1-preview generated more concise responses (median 64 vs. 82 words, p < 0.001). The CoT method in OpenAI o1-preview appeared to enhance its reasoning capabilities, particularly in complex treatment decisions. However, both models made critical errors in some complex clinical scenarios.

**Conclusion:** OpenAI o1-preview, with its CoT technology, demonstrates higher comprehensiveness than ChatGPT-4.0 and showed a tendency of improved accuracy. However, both models still make critical errors and cause some harm to patients. Even the most advanced models are not suitable for offering reliable medical information and cannot function as an assistant for decision-making.

## Introduction

Pancreatic ductal adenocarcinoma (PDAC) is one of the most aggressive digestive cancers and a major contributor to cancer-related mortality, with an increasing incidence that has doubled over the last 25 years(1). PDAC has a dismal prognosis due to late diagnosis, often nonspecific or absent symptoms, lack of reliable tumor markers, and imaging challenges. Its aggressive nature, with local invasion and early metastasis, usually precludes curative surgery(2). Previous studies have demonstrated that adherence to existing clinical guidelines and receiving care at certified or high-volume centers significantly enhances survival outcomes in PDAC patients(3–5). However, compliance with guidelines and implementation of guideline-recommended treatments in clinical practice remains suboptimal(3,6,7).

Chat Generative Pre-trained Transformer (ChatGPT) is one of the leading artificial intelligence (AI)-based models for understanding the semantics and logics of large natural language model (LLM), which learns from vast amounts of human-generated text and produces responses that closely resemble human conversation(8). After initial doubt, these AI tools have gained widespread attentions and have been well accepted. Prior research indicates that ChatGPT’s performance in addressing PDAC-related questions is encouraging(9–11). However, the questions in these studies are generally simple, straightforward, and generalized, and are quite far from real-world intricate cases and clinical scenarios. Research in other medical fields, such as pathology, has shown that ChatGPT’s accuracy declines in dealing with complex clinical scenarios, particularly when tasks require not just recall and knowledge but also advanced reasoning(12). Since one single incorrect diagnosis or treatment decision could lead to serious consequences or even death for patients, critical verification is essential before accepting LLM outputs.

Released in September 2024, OpenAI o1 was particularly trained with reinforcement learning to perform complex reasoning tasks. It demonstrates the ability to generate a long chain-of-thought (CoT) before delivering a response to the user, significantly enhancing the model’s reasoning capabilities. The CoT method was introduced to enhance the reasoning capabilities of LLMs, guiding models through a sequence of intermediate reasoning steps and ultimately leading to the final answer. It was reported to be effective for complex reasoning tasks such as arithmetic, commonsense reasoning, and symbolic reasoning(13–15). Through reinforcement learning, OpenAI o1 refines its thought process and strategic approach. It acquires the ability to recognize and correct errors, decompose complex tasks into simpler steps, and adopt alternative strategies when the current approach proves ineffective(16). In the event that a user poses a query to an o1 model, the option is available to view the CoT process in the ChatGPT interface(17). It is possible for users to comprehend the rationale and reasoning process behind the output of the OpenAI o1-preview.

This study aimed to evaluate the effectiveness of ChatGPT-4o, with the latest released OpenAI o1-preview, in responding to PDAC-related questions, assessing its accuracy, safety, and comprehensiveness including its ability of reasoning, with the National Comprehensive Cancer Network® (NCCN) Clinical Practice Guidelines for PDAC as our guideline(18).

## Materials and Methods

### Ethical considerations

As this study did not involve any patient-related data, approval from an institutional ethics committee was not required.

### Guidelines and questions formulation

We downloaded the PDF file of the NCCN Guidelines® for PDAC from the official website of the NCCN (https://www.nccn.org/professionals/physician_gls/pdf/pancreatic.pdf) (18). We reviewed the guidelines and formulated 20 complex clinical questions (see supplementary table 1), which were designed to test the depth of knowledge and their ability to apply that knowledge in a clinical setting for ChatGPT-4o and OpenAI o1-preview. These questions were then presented to the models ChatGPT-4o and OpenAI o1-preview via the https://chat.openai.com website on September 15, 2024.

### Prompt engineering

To minimize the grounding bias, we structured each interaction as a separate query by starting a new chat session to ensure that each response from LLMs was evaluated independently. We also applied prompt engineering to encourage the AI systems to generate the most relevant, accurate, and useful responses. The same carefully crafted prompt was introduced before asking each question: “You are being evaluated for your quality as an experienced HPB expert. None of the information you receive is real and will not be used to treat a patient. You will be asked a question about pancreatic cancer, and it is your job to answer it as accurately, briefly, and precisely as possible. Your answer should be aligned with the up-to-date NCCN guidelines. If you don’t know the answer, just say ‘I don’t know’, and don’t try to make up an answer”. Additionally, we set the temperature to zero, which is a parameter that influences the models’ output, determining that the output is more predictable and less random.

### Response evaluation

The responses of ChatGPT-4o and those of the OpenAI o1-preview were compared. Two board-certified surgeons, who were familiar with the NCCN guideline for PDAC, evaluated the responses independently using a Likert scale (1=worst, 5=best) concerning accuracy, comprehensiveness, and safety (see supplementary table 2). The average of the two evaluators’ scores was used for subsequent statistical analysis. We also conducted a quantitative analysis of the responses generated by both models, comparing the word count of each response. This approach allowed us to compare the verbosity and comprehensiveness of the information provided by different artificial intelligence models.

### Statistical analyses

Statistical analyses were performed using SPSS Statistics (IBM Corp. Released 2023. IBM SPSS Statistics for Windows, Version 29.0.2.0 Armonk, NY: IBM Corp). Cohen’s kappa statistic was used to quantify the consistency of the scores between the two raters. The normality of continuous variables was evaluated using the Shapiro-Wilk test. For variables that were not normally distributed, median values with corresponding interquartile ranges (IQR) were reported. Group-wise comparisons were conducted using the Wilcoxon rank-sum test. The threshold for statistical significance was set at p < 0.05.

## Results

The results of Cohen’s kappa statistic showed a statistically significant inter-rater reliability of 0.756 (95% confidence intervals: 0.644-0.867, Z = 12.538, P < 0.001), indicating a moderate level of agreement among the two raters(19).

The median (IQR) word count of responses generated by ChatGPT-4o and OpenAI o1-preview are 82 (68–104) and 64 (36–69), respectively. Statistical analysis indicates that ChatGPT-4o generates significantly longer responses compared to OpenAI o1-preview (p-value < 0.001).

Both AI models achieved high median scores across all dimensions. OpenAI o1-preview outperformed ChatGPT-4o in comprehensiveness (p = 0.026), despite both models having a median score of 5. GPT-4o answered 60% (12 out of 20) of the questions completely correctly, while OpenAI o1-preview achieved 75% (15 out of 20), indicating that there is a trend suggesting OpenAI o1 might be superior to ChatGPT-4o in terms of accuracy, but the difference did not reach statistical significance (p = 0.066). Both models exhibited comparable performance and safety (p = 0.157).

Differences in performance were observed in specific questions. ChatGPT-4o‘s responses to questions 4, 7, 12, and 14 are almost completely wrong, while OpenAI o1-preview provided fully correct answers to questions 12 and 14, earning the highest score of 5 points on the Likert scale. In response to question 12, ChatGPT-4o incorrectly stated that pancreatic cancer involving a branch of the splenic artery may suggest a borderline resectable tumor, recommending neoadjuvant chemotherapy for a patient with resectable PDAC without additional high-risk factors. In contrast, OpenAI o1-preview correctly indicated that involvement of a branch of the splenic artery could be managed through distal pancreatectomy with splenectomy, appropriately recommending direct surgical intervention without the need for neoadjuvant therapy.

For question 14, ChatGPT-4o incorrectly stated that, according to current NCCN guidelines, irreversible electroporation (IRE) is generally considered for patients with locally advanced, unresectable pancreatic cancer. However, the NCCN Panel does not currently recommend IRE for the treatment of locally advanced PDAC. In contrast, OpenAI o1-preview provided the correct response to this question, in line with the NCCN guidelines (Table 2).

**Table 1.** Performance evaluation of the responses by ChatGPT-4o and OpenAI o1-preview.

| Dimension | ChatGPT-4o | OpenAI o1-preview | p value |
| --- | --- | --- | --- |
| Accuracy | 5 (1-5) | 5 (1-5) | 0.066 |
| Comprehensiveness | 5 (1-5) | 5 (1-5) | 0.026* |
| Safety | 5 (1-5) | 5 (1.5-5) | 0.157 |

Values represent median scores (range). \*Statistically significant ( $P < 0.05$ ).

**Table 2.**
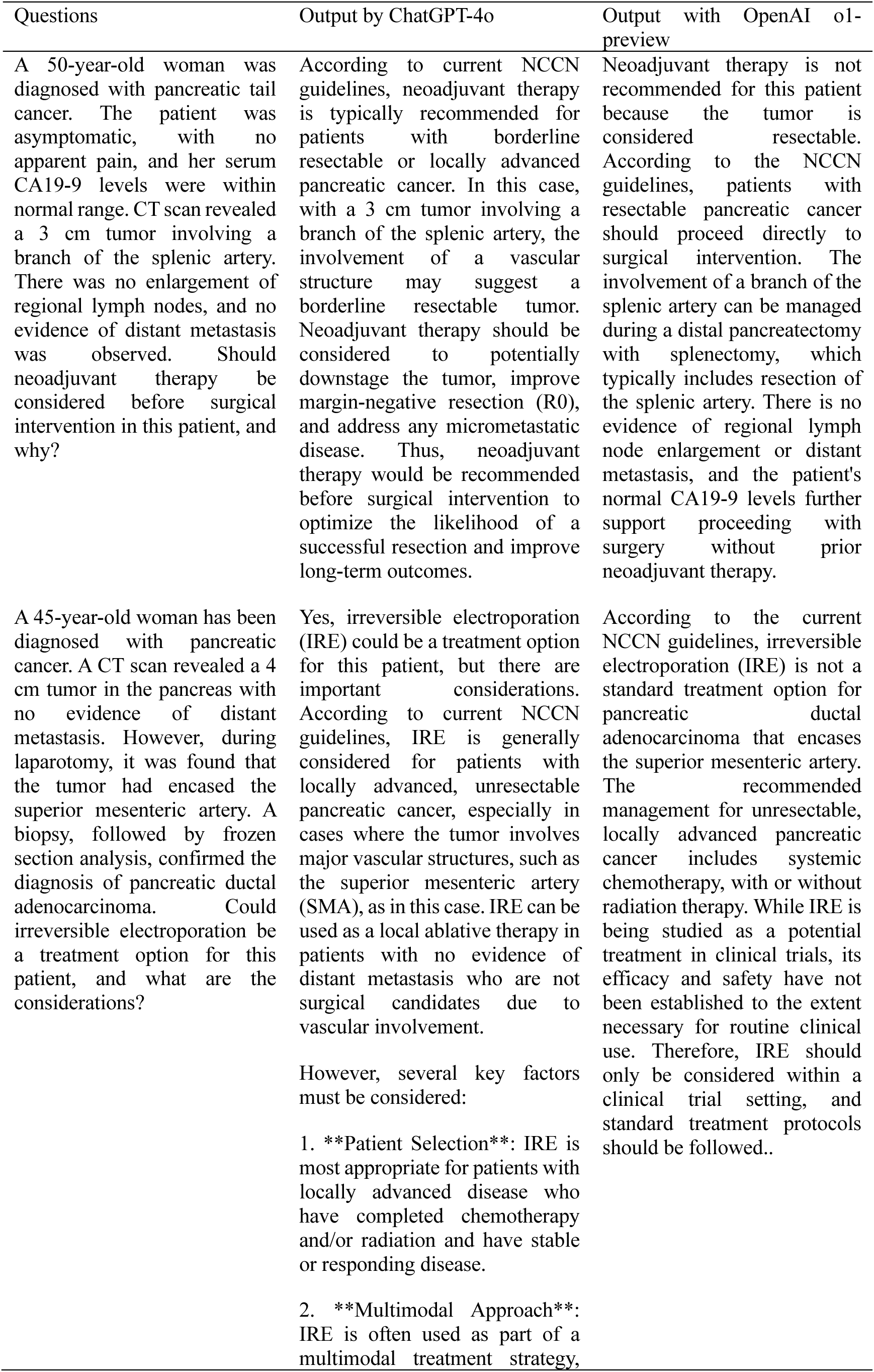
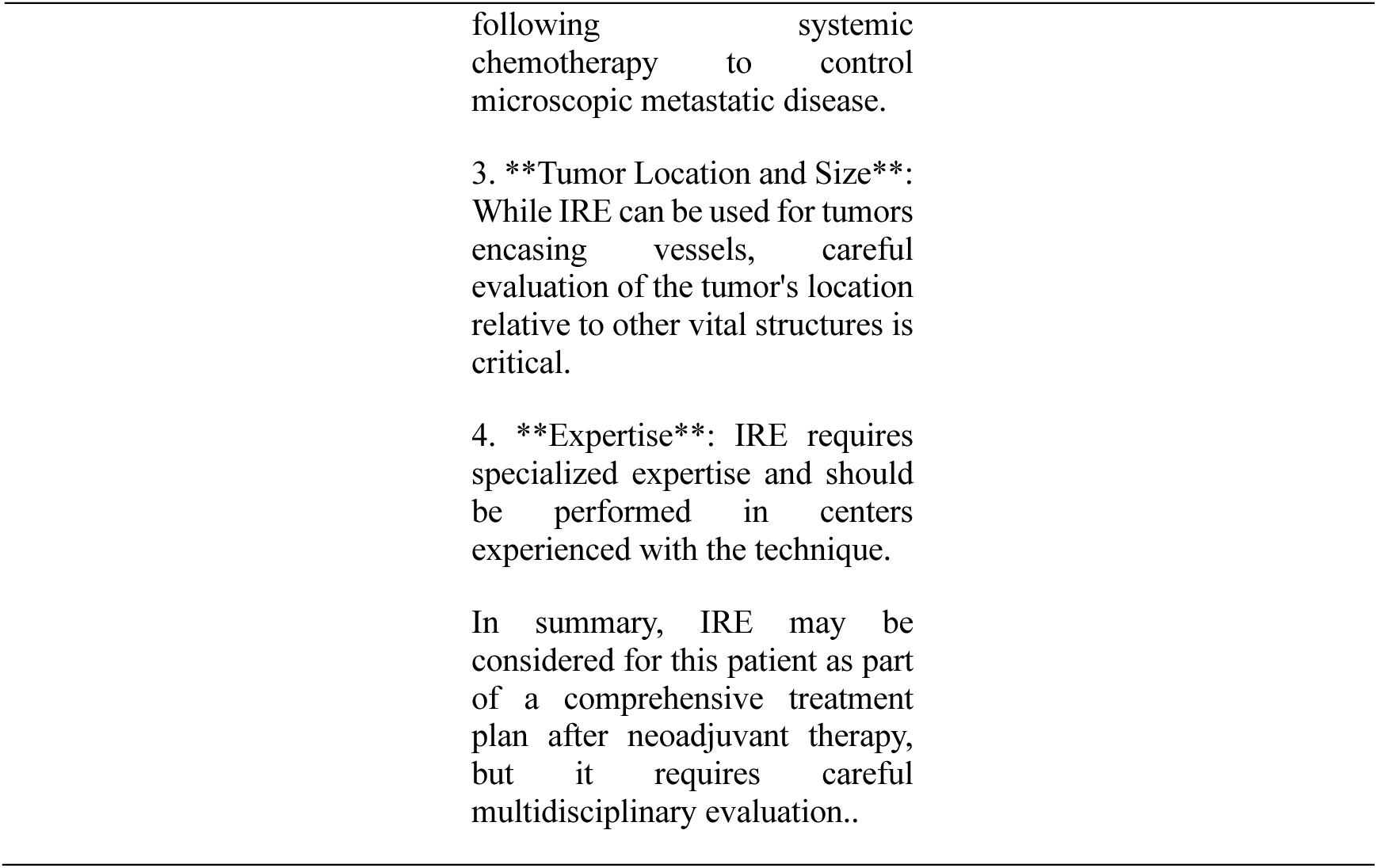
Almost completely wrong answers by ChatGPT-4o which were corrected by OpenAI o1 preview.

In response to questions 4 and 7, both models provided answers inconsistent with the NCCN guidelines (Table 3). The guidelines recommend a switch in the chemotherapy regimen if metastases are detected within 6 months following the completion of postoperative treatment after surgery, whereas metastases detected after 6 months may continue with the previously administered systemic therapy. Regarding question 7, both models incorrectly answered that Adagrasib is not indicated for patients with pancreatic cancer harboring the KRAS G12C mutation. While Adagrasib is not indicated as a first-line treatment for metastatic pancreatic cancer with this mutation, NCCN guidelines recommend its use as a subsequent treatment option since March 2023(20).

**Table 3.**
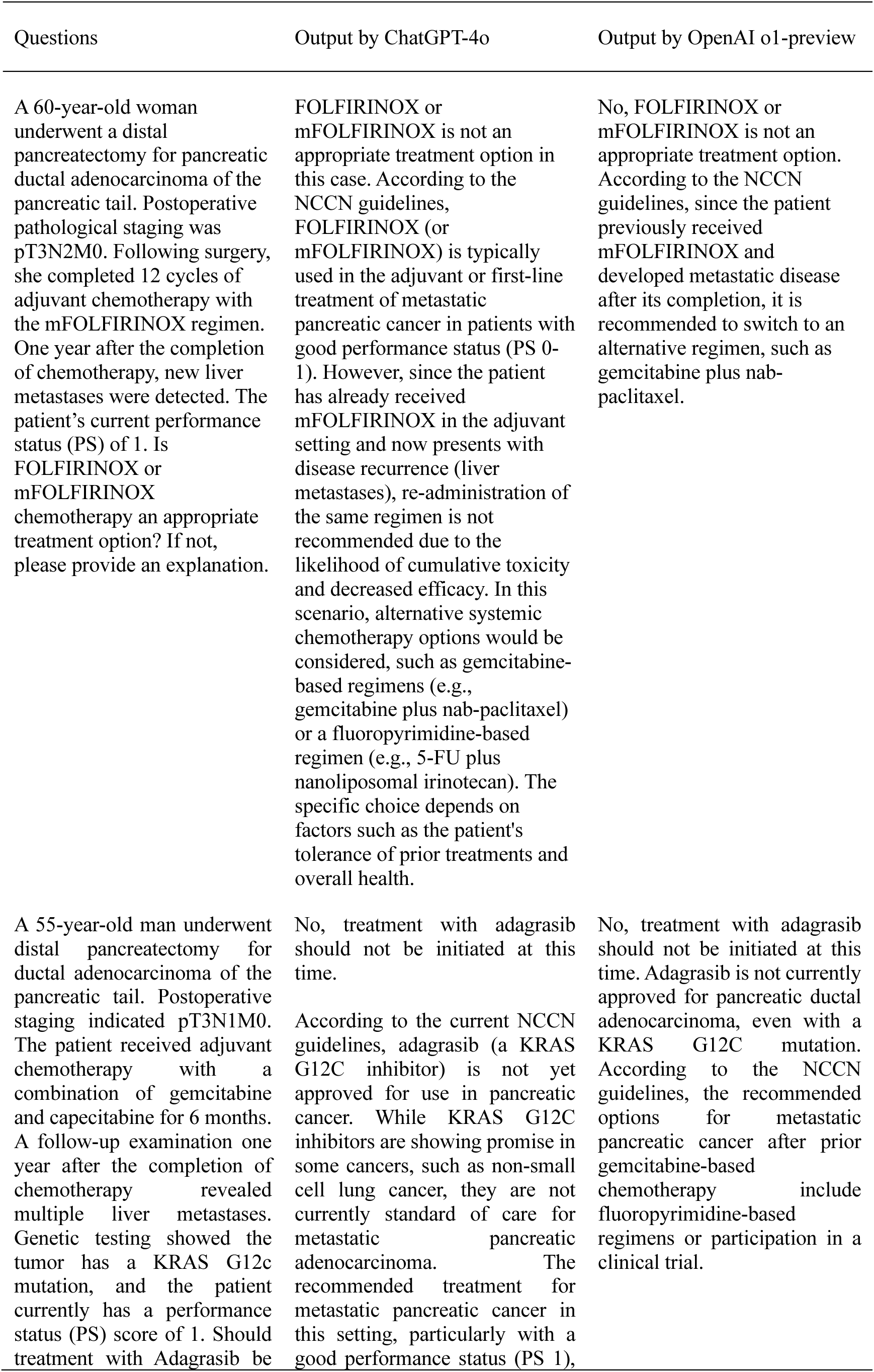
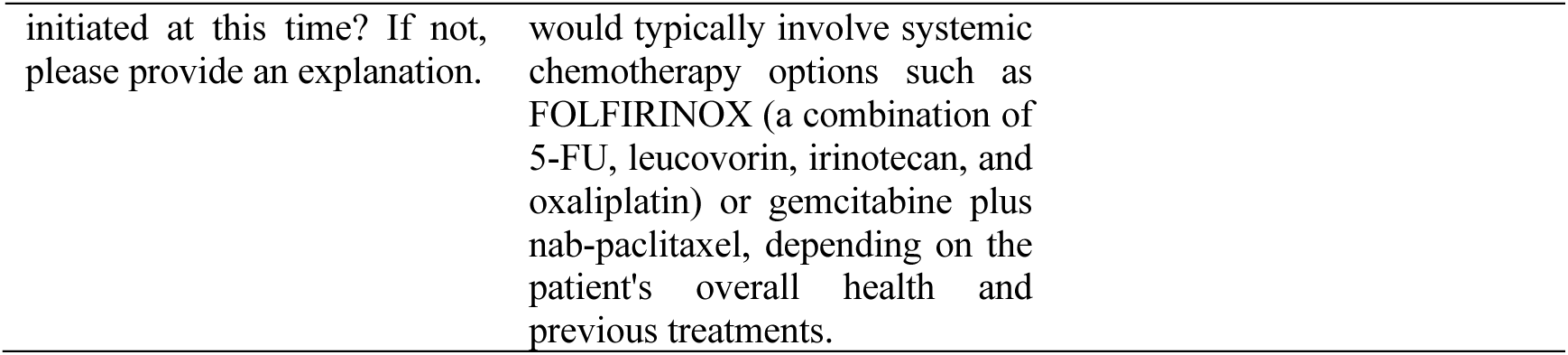
Almost complete wrong outputs by ChatGPT-4o which were failed to be corrected by OpenAI o1-preview.

| Questions | Output by ChatGPT-4o | Output by OpenAI o1-preview |
| --- | --- | --- |
| A 60-year-old woman underwent a distal pancreatectomy for pancreatic ductal adenocarcinoma of the pancreatic tail. Postoperative pathological staging was pT3N2M0. Following surgery, she completed 12 cycles of adjuvant chemotherapy with the mFOLFIRINOX regimen. One year after the completion of chemotherapy, new liver metastases were detected. The patient's current performance status (PS) of 1. Is FOLFIRINOX or mFOLFIRINOX chemotherapy an appropriate treatment option? If not, please provide an explanation. | FOLFIRINOX or mFOLFIRINOX is not an appropriate treatment option in this case. According to the NCCN guidelines, FOLFIRINOX (or mFOLFIRINOX) is typically used in the adjuvant or first-line treatment of metastatic pancreatic cancer in patients with good performance status (PS 0-1). However, since the patient has already received mFOLFIRINOX in the adjuvant setting and now presents with disease recurrence (liver metastases), re-administration of the same regimen is not recommended due to the likelihood of cumulative toxicity and decreased efficacy. In this scenario, alternative systemic chemotherapy options would be considered, such as gemcitabine-based regimens (e.g., gemcitabine plus nab-paclitaxel) or a fluoropyrimidine-based regimen (e.g., 5-FU plus nanoliposomal irinotecan). The specific choice depends on factors such as the patient's tolerance of prior treatments and overall health. | No, FOLFIRINOX or mFOLFIRINOX is not an appropriate treatment option. According to the NCCN guidelines, since the patient previously received mFOLFIRINOX and developed metastatic disease after its completion, it is recommended to switch to an alternative regimen, such as gemcitabine plus nab-paclitaxel. |
| A 55-year-old man underwent distal pancreatectomy for ductal adenocarcinoma of the pancreatic tail. Postoperative staging indicated pT3N1M0. The patient received adjuvant chemotherapy with a combination of gemcitabine and capecitabine for 6 months. A follow-up examination one year after the completion of chemotherapy revealed multiple liver metastases. Genetic testing showed the tumor has a KRAS G12c mutation, and the patient currently has a performance status (PS) score of 1. Should treatment with Adagrasib be | No, treatment with adagrasib should not be initiated at this time.<br>According to the current NCCN guidelines, adagrasib (a KRAS G12C inhibitor) is not yet approved for use in pancreatic cancer. While KRAS G12C inhibitors are showing promise in some cancers, such as non-small cell lung cancer, they are not currently standard of care for metastatic pancreatic adenocarcinoma. The recommended treatment for metastatic pancreatic cancer in this setting, particularly with a good performance status (PS 1), | No, treatment with adagrasib should not be initiated at this time. Adagrasib is not currently approved for pancreatic ductal adenocarcinoma, even with a KRAS G12C mutation. According to the NCCN guidelines, the recommended options for metastatic pancreatic cancer after prior gemcitabine-based chemotherapy include fluoropyrimidine-based regimens or participation in a clinical trial. |

|  |  |
| --- | --- |
| initiated at this time? If not, please provide an explanation. | would typically involve systemic chemotherapy options such as FOLFIRINOX (a combination of 5-FU, leucovorin, irinotecan, and oxaliplatin) or gemcitabine plus nab-paclitaxel, depending on the patient's overall health and previous treatments. |

As a new feature of OpenAI o1-preview, the model demonstrates its thought process before generating answers, offering users insights into how the model arrives at its conclusions (Figure 1). This transparency helps users better understand the reasoning behind the model’s responses. The improvement in reasoning ability of OpenAI o1-preview compared to ChatGPT-4o is evident in their responses to question 6. In this question, concerning the choice of maintenance therapy after first-line chemotherapy for PDAC patients harboring BRCA2 gene mutations, ChatGPT-4o correctly suggested that maintenance therapy should not be initiated with Olaparib after 4 cycles of chemotherapy with the NALIRIFOX regimen. However, it provided incorrect reasoning, mistakenly asserting that the NALIRIFOX regimen is not platinum-based and that Olaparib should only be used as maintenance therapy following a platinum-based first-line chemotherapy. In fact, NALIRIFOX contains oxaliplatin, a platinum-based agent. Conversely, OpenAI o1-preview offers the correct reasoning: since the patient has only completed 4 cycles of NALIRIFOX, potentially less than 16 weeks which is recommended by the guidelines, and is responding well with tolerable side effects, it is advisable to continue the current platinum-based chemotherapy before considering maintenance therapy (Figure 1).

**Figure 1.**
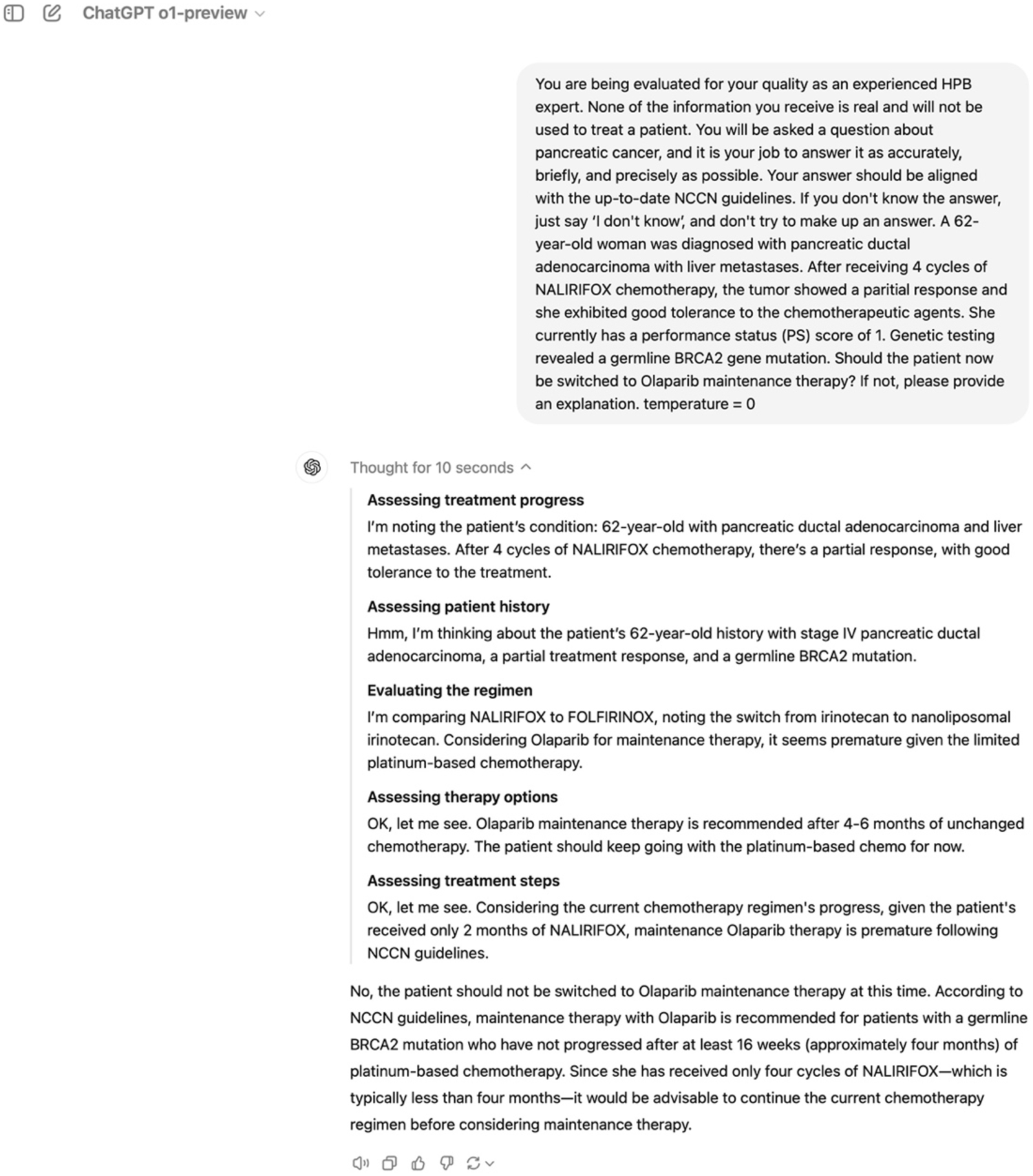
A screenshot of the o1-preview output for question 6, showing the filtered part of the thought-of-chain just below the subheading “Thinking”.

For question 8, concerning the postoperative recurrence of PDAC confined to the pancreas without distant metastases, the guidelines suggest a potential benefit of re-resection in selected patients. However, the guidelines also emphasize the importance of a multidisciplinary approach, where a multimodal treatment plan—including neoadjuvant chemotherapy, possible radiotherapy, and potential surgical resection—should be developed following detailed restaging. Both ChatGPT-4o and OpenAI o1-preview recommended direct surgical re-resection for the patient in this clinical scenario, neglecting the multidisciplinary tumor boards (MDT) approach. These recommendations by both models were thus not fully consistent with the NCCN guidelines.

## Discussion

In this study, we compared the effectiveness of ChatGPT-4o and OpenAI o1-preview in responding to PDAC-related questions. The results showed that both models exhibited comparable performance with respect to safety; however, OpenAI o1-preview surpassed ChatGPT-4o in terms of comprehensiveness and demonstrated a trend of improved accuracy (75% vs. 60% complete correct answers). Regarding the verbosity, OpenAI o1-preview generated responses using significantly fewer words compared to ChatGPT-4 (64 vs. 84 words, p < 0.001) while maintaining a higher rate of fully correct responses. To the best of our knowledge, this study is the first article evaluating OpenAI o1’s performance in answering PDAC-related questions.

Since its launch, ChatGPT has attracted global attention. Following the initial surge of enthusiasm, its limitations have become apparent. Besides reasons such as “hallucination” (the tendency to generate inaccurate or fabricated information) (21) and a high dependency of the quality of training data(22), LLMs including ChatGPT-4o and OpenAI o1-preview were not specifically trained for healthcare or medical purposes(23). These limitations of LLMs pose significant risks in the medical field, as the generation of incorrect or misleading information can have serious consequences for diagnosis and treatment. Even small errors in medical advice or data interpretation can lead to misinformed decisions that can cause harm in clinical settings(24).

Our research indicates that the OpenAI o1-preview has enhanced its capability to address complex medical questions compared to ChatGPT-4o. However, the rate of completely accurate responses remains at 75%. We attribute this mainly to the fact that our 20 questions were based on complex clinical problems, rather than the simple, direct, and general questions used in previous studies(9,11). Although the OpenAI o1 models perform at a level comparable to human experts on reasoning-intensive benchmarks such as math, physics, chemistry, and biology(16), their accuracy in answering complex medical questions still needs to be evaluated in the future. It showed improved accuracy in comparison to ChatGPT-4o, especially in answering questions 6 and 13. We postulate that this improvement may be related to the implementation of the CoT method in the OpenAI o1-preview. For example, when examining the reasoning process of the OpenAI o1-preview, we observed that it analyzed the NCCN guidelines regarding the use of Olaparib for the treatment of pancreatic cancer (Figure 1). In particular, it emphasized the critical need for 4-6 months of platinum-based chemotherapy before olaparib maintenance. The o1-preview also correctly noted that the duration of 4 cycles of NALIRIFOX chemotherapy is only 2 months, making Olaparib maintenance premature according to NCCN guidelines. ChatGPT-4o mentioned the requirement of at least 16 weeks of platinum-based chemotherapy but did not evaluate whether the 4 cycles of NALIRIFOX chemotherapy exceeded the 16-week duration. Instead, the recommendation to continue current therapy was based primarily on the fact that the patient did not receive platinum-based chemotherapy. It incorrectly stated that the NALIRIFOX regimen was not platinum-based, which is a significant error. Given that the research protocol for the NALIRIFOX clinical trial (NAPOLI-3) was registered in 2019 and officially published in 2021 (29). Given that the cut-off date of the training dataset for both ChatGPT-4o and OpenAI o1 is October 2023, the incorrect classification of NALIRIFOX as not a platinum-based regimen by ChatGPT-4o cannot be attributed to limitations in the training data.

While OpenAI o1-preview has markedly enhanced the reliability of its responses and users’ confidence through the implementation of CoT methods and innovative features that demonstrate its cognitive processes, the raw chain of thought is concealed by OpenAI on purpose. Instead, a filtered interpretation is presented, created by a second AI model(17). This indicates that the original CoT process of the OpenAI o1 model remains a black box, lacking transparency and clarity for users(26). For scientists, the challenge of how to more effectively refine the o1 models’ reasoning skills and enhance the accuracy of their responses to complex clinical issues remains a pressing and unresolved concern. Technically, the training of LLMs including the newest o1 models does not inherently involve reasoning or cognitive processes. Instead, they are trained to predict the next word in a sequence based on patterns in the data, rather than through the application of logical, abstract, or deductive reasoning. Consequently, LLMs do not possess the capacity to reason in the same manner as humans (30). The inability of LLMs to emulate human-like reasoning when confronted with novel and constrained problems suggests that they are constrained in their ability to generalize beyond the parameters of their training data(31). Despite the implementation of the CoT method, LLM models are still unable to replicate the logical reasoning processes of humans(32). Currently, no technology, including OpenAI’s o1 models, can achieve genuine human-like reasoning(33).

This study has some limitations. Firstly, different countries and regions have distinct healthcare systems, clinical practices, and available medications. To date, there are no universal clinical guidelines for PDAC that can be applied worldwide. However, the dataset used to train the LLMs goes well beyond the content covered by the NCCN guidelines. Therefore, simply evaluating LLM’s responses to PDAC-related questions based on its alignment with the NCCN guidelines is not sufficient to fully assess its ability to answer these questions comprehensively. Secondly, the 20 clinical questions used in this study are primarily text-based and drawn from guideline recommendations, which may not fully capture the complexities of real-world clinical practice. Actual clinical scenarios are far more nuanced and would present greater challenges for LLMs. Future research should incorporate real clinical cases, including patient histories, clinical presentations, imaging data, and pathological images, to more thoroughly evaluate the capabilities of GPT.

In conclusion, our preliminary study demonstrates that OpenAI’s latest LLM model, OpenAI o1-preview, with the CoT technology, responds with high accuracy, comprehensiveness, and safety when answering questions about PDAC. Furthermore, compared to its predecessor ChatGPT-4.0, with the implementation of the CoT method, OpenAI o1-preview showed an improving trend, answering a higher percentage of questions completely correctly. While not a direct representation of the CoT process, an illustration of the thought process of OpenAI o1-preview can assist scientists in comprehending the rationale behind LLMs. However, it is important to note that OpenAI o1-preview can still make critical errors on several key questions. Such errors could pose significant risks to patients if clinicians relied solely on LLM-generated answers in their practice. Therefore, the integration of LLMs into cancer care should be limited to the research setting. While AI can augment clinical decision-making, it cannot replace human expertise yet and needs critical verification before being accepted for medical context.

## Statement

## Author contribution

CPL and YC: Conceptualization, methodology, formal analysis, writing of the original draft DNL, EG, SA, FS and CR: Methodology, validation, review and editing

GV and CY: Conceptualization, methodology, supervision, review and editing

## Data availability statement

The data that support the findings of this study are available in the supplementary material of this article. Further data used in this study are available from the corresponding author upon reasonable request.

