## Supplementary Table 1 for "Close, But no Cigar: Comparative Evaluation of ChatGPT-4o and OpenAI o1-preview in Answering Pancreatic Ductal Adenocarcinoma-Related Questions"

**Supplementary table 1** 20 complex clinical questions

| No. | Question | Answers by ChatGPT-4o | Answers by OpenAI o1 |
| --- | --- | --- | --- |
| 1 | A 50-year-old male patient diagnosed with pancreatic ductal adenocarcinoma and liver metastasis, then he underwent six cycles of chemotherapy. Remarkably, both the baseline serum CA19-9 levels and those measured during chemotherapy were within the normal range. What is the most likely explanation for the patient's normal CA19-9? | The most likely explanation for the patient's normal CA19-9 levels, despite having pancreatic ductal adenocarcinoma with liver metastasis, is that the patient is a Lewis antigen-negative individual (Lea-b-). About 5-10% of the population lacks the Lewis antigen and, as a result, cannot produce CA19-9, leading to normal or undetectable levels even in the presence of pancreatic cancer. | The most likely explanation is that the patient is Lewis antigen-negative (Le⁻a⁻b⁻) and therefore cannot produce CA19-9, leading to normal serum levels despite having pancreatic cancer. |
| 2 | A 45-year-old man presented with pancreatic ductal adenocarcinoma of the uncinate process. Contrast CT imaging revealed no metastatic lesions outside the pancreas. The tumor contacted more than 180° of the portal vein, but the involved portion of the vein could be safely resected and reconstructed. Additionally, the tumor fully encircled the superior mesenteric artery. Considering these findings, would this patient be classified as having borderline resectable pancreatic cancer? If not, please provide an explanation. | This patient would **not** be classified as having borderline resectable pancreatic cancer. According to the **NCCN guidelines**, the key factor here is the **superior mesenteric artery (SMA) encasement**. Tumors that fully encircle the SMA are considered **unresectable** rather than borderline resectable. While the portal vein involvement with the potential for safe resection and reconstruction aligns with criteria for borderline resectable, the extent of SMA involvement precludes this classification. Therefore, the tumor is considered **locally advanced/unresectable**. | No, this patient would not be classified as having borderline resectable pancreatic cancer. According to the NCCN guidelines, tumor encasement of the superior mesenteric artery (SMA) greater than 180° (i.e., fully encircling the artery) is considered unresectable disease. Therefore, the full encasement of the SMA by the tumor places this patient in the unresectable category. |
| 3 | A 42-year-old man underwent a pancreaticoduodenectomy for pancreatic ductal adenocarcinoma. Postoperative pathological staging revealed pT2N1M0. Following surgery, he completed 12 cycles of adjuvant chemotherapy with the mFOLFIRINOX regimen. Three months after completing chemotherapy, new liver metastases were detected. Given that the patient currently has a performance status (PS) score of 1, is fluoropyrimidine-based systemic chemotherapy an appropriate treatment option? If not, please provide an explanation. | No, fluoropyrimidine-based systemic chemotherapy alone would not be the most appropriate treatment option in this case. According to the NCCN guidelines, in a patient with recurrent metastatic pancreatic cancer after receiving adjuvant FOLFIRINOX, second-line treatment options typically include gemcitabine-based therapy (e.g., gemcitabine plus nab-paclitaxel) or consideration of clinical trials. Given that the patient has a performance status of 1, he would likely be eligible for further systemic therapy, but a fluoropyrimidine-based regimen would not be recommended as the primary option after progression following FOLFIRINOX. | No, fluoropyrimidine-based systemic chemotherapy is not appropriate in this case. The patient has already been treated with mFOLFIRINOX, which includes 5-fluorouracil—a fluoropyrimidine. According to the NCCN guidelines, patients who progress after fluoropyrimidine-based therapy should be considered for alternative treatments like gemcitabine plus nab-paclitaxel. |
| 4 | A 60-year-old woman underwent a distal pancreatectomy for pancreatic ductal adenocarcinoma of the pancreatic tail. Postoperative pathological staging was pT3N2M0. Following surgery, she completed 12 cycles of adjuvant chemotherapy with the mFOLFIRINOX regimen. One year after the completion of chemotherapy, new liver metastases were detected. The patient’s current performance status (PS) of 1. Is FOLFIRINOX or mFOLFIRINOX chemotherapy an appropriate treatment option? If not, please provide an explanation. | FOLFIRINOX or mFOLFIRINOX is not an appropriate treatment option in this case. According to the NCCN guidelines, FOLFIRINOX (or mFOLFIRINOX) is typically used in the adjuvant or first-line treatment of metastatic pancreatic cancer in patients with good performance status (PS 0-1). However, since the patient has already received mFOLFIRINOX in the adjuvant setting and now presents with disease recurrence (liver metastases), re-administration of the same regimen is not recommended due to the likelihood of cumulative toxicity and decreased efficacy.  In this scenario, alternative systemic chemotherapy options would be considered, such as gemcitabine-based regimens (e.g., gemcitabine plus nab-paclitaxel) or a fluoropyrimidine-based regimen (e.g., 5-FU plus nanoliposomal irinotecan). The specific choice depends on factors such as the patient's tolerance of prior treatments and overall health. | No, FOLFIRINOX or mFOLFIRINOX is not an appropriate treatment option. According to the NCCN guidelines, since the patient previously received mFOLFIRINOX and developed metastatic disease after its completion, it is recommended to switch to an alternative regimen, such as gemcitabine plus nab-paclitaxel. |
| 5 | A 53-year-old man was diagnosed with resectable pancreatic cancer, with the tumor located in the head of the pancreas. CT revealed significant dilatation of the intra- and extrahepatic bile ducts, along with an enlarged gallbladder. His total serum bilirubin level is 180 µmol/L. The patient is scheduled for surgery in 5 days but now presents with abdominal pain, fever, and elevated leukocyte and CRP levels. Should the patient be considered for biliary stenting via ERCP? | Yes, the patient should be considered for biliary stenting via ERCP. According to NCCN guidelines, patients with resectable pancreatic cancer who present with obstructive jaundice, particularly those with significant symptoms like fever, abdominal pain, and signs of cholangitis (as evidenced by elevated leukocyte and CRP levels), should undergo biliary decompression before surgery. ERCP with stent placement is recommended in this scenario to manage biliary obstruction and reduce the risk of further complications such as cholangitis before the planned surgery. | Yes, the patient should undergo biliary stenting via ERCP to manage acute cholangitis before surgery. According to the NCCN guidelines, preoperative biliary drainage is recommended in cases of cholangitis to relieve obstruction and treat the infection. |
| 6 | A 62-year-old woman was diagnosed with pancreatic ductal adenocarcinoma with liver metastases. After receiving 4 cycles of NALIRIFOX chemotherapy, the tumor showed a paritial response and she exhibited good tolerance to the chemotherapeutic agents. She currently has a performance status (PS) score of 1. Genetic testing revealed a germline BRCA2 gene mutation. Should the patient now be switched to olaparib maintenance therapy? If not, please provide an explanation. | No, the patient should not be switched to olaparib maintenance therapy at this time. According to the NCCN guidelines, olaparib maintenance therapy is recommended for patients with germline BRCA1/2 mutations who have received at least 16 weeks (or 4 months) of a platinum-based chemotherapy regimen and achieved either a complete or partial response. The NALIRIFOX regimen includes irinotecan, which is not platinum-based. Therefore, the patient would not be eligible for olaparib maintenance therapy at this stage unless she had been treated with a platinum-based regimen like FOLFIRINOX. | No, the patient should not be switched to Olaparib maintenance therapy at this time. According to NCCN guidelines, maintenance therapy with Olaparib is recommended for patients with a germline BRCA2 mutation who have not progressed after at least 16 weeks (approximately four months) of platinum-based chemotherapy. Since she has received only four cycles of NALIRIFOX-which is typically less than four months-it would be advisable to continue the current chemotherapy regimen before considering maintenance therapy. |
| 7 | A 55-year-old man underwent distal pancreatectomy for ductal adenocarcinoma of the pancreatic tail. Postoperative staging indicated pT3N1M0. The patient received adjuvant chemotherapy with a combination of gemcitabine and capecitabine for 6 months. A follow-up examination one year after the completion of chemotherapy revealed multiple liver metastases. Genetic testing showed the tumor has a KRAS G12c mutation, and the patient currently has a performance status (PS) score of 1. Should treatment with Adagrasib be initiated at this time? If not, please provide an explanation. | No, treatment with adagrasib should not be initiated at this time.   According to the current NCCN guidelines, adagrasib (a KRAS G12C inhibitor) is not yet approved for use in pancreatic cancer. While KRAS G12C inhibitors are showing promise in some cancers, such as non-small cell lung cancer, they are not currently standard of care for metastatic pancreatic adenocarcinoma. The recommended treatment for metastatic pancreatic cancer in this setting, particularly with a good performance status (PS 1), would typically involve systemic chemotherapy options such as FOLFIRINOX (a combination of 5-FU, leucovorin, irinotecan, and oxaliplatin) or gemcitabine plus nab-paclitaxel, depending on the patient's overall health and previous treatments. | No, treatment with adagrasib should not be initiated at this time. Adagrasib is not currently approved for pancreatic ductal adenocarcinoma, even with a KRAS G12C mutation. According to the NCCN guidelines, the recommended options for metastatic pancreatic cancer after prior gemcitabine-based chemotherapy include fluoropyrimidine-based regimens or participation in a clinical trial. |
| 8 | A 40-year-old male underwent distal pancreatectomy for ductal adenocarcinoma of the pancreatic tail. Postoperative staging was pT3N0M0. The patient received chemotherapy with the mFOLFIRINOX regimen for 6 months following surgery. A follow-up conducted 6 months after the completion of chemotherapy revealed a newly detected local recurrence at the pancreatic margin, measuring approximately 1 cm in diameter. Systemic staging showed no distant metastases, with the recurrence limited to the local site. The surgeons evaluated the recurrence as resectable. What would be the appropriate treatment strategy for this patient, and please explain the rationale for it? | The appropriate treatment strategy for this patient, according to up-to-date NCCN guidelines, would be **surgical re-resection** of the local recurrence, followed by **consideration of additional chemotherapy or chemoradiation therapy**.  **Rationale**: 1. **Surgical resection** is the mainstay treatment for localized, resectable recurrence of pancreatic adenocarcinoma. Since the recurrence is confined to the local site and evaluated as resectable, surgery offers the best chance for disease control and potential long-term survival.  2. **Post-resection therapy**: After surgical re-resection, consideration should be given to **adjuvant chemotherapy** or **chemoradiation**, depending on the patient's performance status, previous treatment response, and other clinical factors. Given that the patient has already received mFOLFIRINOX, reintroduction of chemotherapy or transition to chemoradiation (if not previously used) could be considered to maximize local control and minimize the risk of further recurrence.  In summary, the patient should undergo **surgical re-resection** followed by evaluation for **adjuvant therapy**. | The appropriate treatment strategy is to proceed with surgical resection of the local recurrence (re-resection). This is followed by consideration of additional systemic therapy or enrollment in a clinical trial.  **Rationale:**  - **Resectable Local Recurrence:** The patient has a resectable local recurrence without distant metastases. Surgical resection offers the best chance for local control and potential long-term survival in such cases.   - **Previous Treatment History:** Given that the patient has already received adjuvant chemotherapy with mFOLFIRINOX, re-resection provides an opportunity to manage the disease that has recurred despite systemic therapy.   - **Young Age and Good Performance Status:** At 40 years old, the patient is likely to tolerate additional surgery and postoperative therapies well.   - **Guideline Alignment:** According to up-to-date NCCN guidelines, re-resection can be considered for isolated local recurrences in select patients to improve outcomes. |
| 9 | A 57-year-old man underwent a pancreaticoduodenectomy for obstructive jaundice and a pancreatic head mass. Postoperative pathology confirmed adenosquamous carcinoma, with staging pT2N2M0. Should this patient's postoperative follow-up and adjuvant therapy follow the same strategy as that for pancreatic ductal adenocarcinoma? | Yes, the postoperative follow-up and adjuvant therapy for this patient should follow the same strategy as that for pancreatic ductal adenocarcinoma (PDAC). According to current NCCN guidelines, adenosquamous carcinoma of the pancreas, despite its unique histology, is treated similarly to PDAC due to its aggressive nature and poor prognosis. This typically includes adjuvant chemotherapy, most commonly with gemcitabine-based or fluoropyrimidine-based regimens, and may involve radiation therapy in certain cases. Follow-up should include regular clinical assessments, CA 19-9 tumor marker monitoring, and imaging as per the standard surveillance for PDAC. | Yes, according to current NCCN guidelines, the postoperative follow-up and adjuvant therapy for adenosquamous carcinoma of the pancreas should follow the same strategy as that for pancreatic ductal adenocarcinoma. |
| 10 | A 45-year-old man presented with a pancreatic body tumor. A CT scan revealed a 2.5 × 3 cm hypovascular mass, with no evidence of enlarged regional lymph nodes or distant metastases. The preoperative diagnosis was resectable pancreatic cancer. Folloing a MDT discussion, laparoscopic distal pancreatectomy was recommended. During laparoscopic exploration, a 0.5 cm grayish-white nodule was identified in the greater omentum. The nodule was resected, and frozen section pathology confirmed it as a metastasis of ductal adenocarcinoma. Should the laparoscopic distal pancreatectomy proceed? Please provide an explanation. | No, the laparoscopic distal pancreatectomy should not proceed. The identification of a metastatic nodule in the greater omentum confirms stage IV pancreatic ductal adenocarcinoma, according to the NCCN guidelines. Surgery with curative intent is not indicated in the presence of metastatic disease. Instead, systemic therapy should be considered as the primary treatment approach for metastatic pancreatic cancer. | No, the laparoscopic distal pancreatectomy should not proceed. The discovery of a metastatic nodule in the greater omentum confirms stage IV disease with distant metastasis. According to the NCCN guidelines, surgical resection is not recommended for metastatic pancreatic cancer. The appropriate course of action is to initiate systemic chemotherapy. |
| 11 | A 40-year-old man was admitted to the hospital with abdominal pain, and a mass was found in the neck of the pancreas. CT imaging showed that the tumor encased the celiac artery, with no lymph node enlargement or distant metastasis. The patient underwent an endoscopic ultrasound-guided biopsy, which revealed pancreatic ductal adenocarcinoma. Genetic testing indicated high microsatellite instability (MSI-H) in the tumor, with a tumor mutational burden (TMB) of 12 mut/mb. The patient also exhibited a KRAS 12c mutation and a P53 mutation, with no other gene mutations detected. The patient currently has a performance status (PS) score of 2. Is treatment with Durvalumab appropriate for the patient at present? Please provide an explanation. | No, treatment with Durvalumab is not currently appropriate for this patient based on the NCCN guidelines. While high microsatellite instability (MSI-H) suggests that immunotherapy could be considered, Durvalumab is not the standard immunotherapy agent for MSI-H pancreatic cancer. According to the NCCN guidelines, Pembrolizumab is the recommended immune checkpoint inhibitor for patients with MSI-H or mismatch repair deficient (dMMR) pancreatic adenocarcinoma. Additionally, Durvalumab is not currently listed as a treatment option for pancreatic cancer in the guidelines.  Therefore, if immunotherapy is considered due to the MSI-H status, Pembrolizumab would be the preferred option. Further evaluation of the patient's overall condition, including their performance status (PS 2), is important in making a treatment decision, as PS may influence the choice and timing of systemic therapy. | No, treatment with Durvalumab is not appropriate for this patient at present. According to the NCCN guidelines, pembrolizumab is the recommended immunotherapy for pancreatic ductal adenocarcinoma with high microsatellite instability (MSI-H) or deficient mismatch repair (dMMR). Durvalumab is not currently recommended for this indication in pancreatic cancer. |
| 12 | A 50-year-old woman was diagnosed with pancreatic tail cancer. The patient was asymptomatic, with no apparent pain, and her serum CA19-9 levels were within normal range. CT scan revealed a 3 cm tumor involving a branch of the splenic artery. There was no enlargement of regional lymph nodes, and no evidence of distant metastasis was observed. Should neoadjuvant therapy be considered before surgical intervention in this patient, and why? | According to current NCCN guidelines, neoadjuvant therapy is typically recommended for patients with borderline resectable or locally advanced pancreatic cancer. In this case, with a 3 cm tumor involving a branch of the splenic artery, the involvement of a vascular structure may suggest a borderline resectable tumor. Neoadjuvant therapy should be considered to potentially downstage the tumor, improve margin-negative resection (R0), and address any micrometastatic disease.  Thus, neoadjuvant therapy would be recommended before surgical intervention to optimize the likelihood of a successful resection and improve long-term outcomes. | Neoadjuvant therapy is not recommended for this patient because the tumor is considered resectable. According to the NCCN guidelines, patients with resectable pancreatic cancer should proceed directly to surgical intervention. The involvement of a branch of the splenic artery can be managed during a distal pancreatectomy with splenectomy, which typically includes resection of the splenic artery. There is no evidence of regional lymph node enlargement or distant metastasis, and the patient's normal CA19-9 levels further support proceeding with surgery without prior neoadjuvant therapy. |
| 13 | A 53-year-old man was referred due to abdominal pain. A CT scan revealed a 3.5 cm tumor in the tail of the pancreas, along with several lymph nodes in the splenic hilum showing a short diameter greater than 2 cm. No distant metastases were identified. The CA19-9 level was 610 U/mL at the initial outpatient visit two weeks ago, and upon rechecking at this visit, it had increased to 1105 U/mL. Should neoadjuvant therapy be considered before surgical intervention in this patient, and why? | Yes, neoadjuvant therapy should be considered before surgical intervention in this patient. According to the NCCN guidelines for pancreatic cancer, patients with radiographically suspicious regional lymph nodes (such as those in the splenic hilum greater than 2 cm) and elevated CA19-9 levels (especially with a significant increase) are at high risk for micrometastatic disease and poor outcomes after upfront surgery. Neoadjuvant therapy can help downstage the tumor, improve resectability, address potential micrometastatic disease, and provide an early assessment of tumor biology and response to treatment.  Therefore, neoadjuvant therapy is recommended to optimize outcomes before considering surgical resection. | Yes, neoadjuvant therapy should be considered before surgical intervention in this patient. The reasons are:  - **High-Risk Features**: The presence of enlarged lymph nodes (>2 cm in the splenic hilum) and a rapidly rising CA19-9 level indicate aggressive tumor biology.   - **Potential Micrometastatic Disease**: Elevated tumor markers suggest the possibility of micrometastatic disease, which neoadjuvant therapy can address.   - **Improving Surgical Outcomes**: Neoadjuvant therapy may downstage the tumor and involved lymph nodes, increasing the likelihood of a successful resection and better long-term outcomes.  According to the NCCN guidelines, patients with such high-risk features benefit from systemic therapy prior to surgery to improve survival rates and reduce recurrence. |
| 14 | A 45-year-old woman has been diagnosed with pancreatic cancer. A CT scan revealed a 4 cm tumor in the pancreas with no evidence of distant metastasis. However, during laparotomy, it was found that the tumor had encased the superior mesenteric artery. A biopsy, followed by frozen section analysis, confirmed the diagnosis of pancreatic ductal adenocarcinoma. Could irreversible electroporation be a treatment option for this patient, and what are the considerations? | Yes, irreversible electroporation (IRE) could be a treatment option for this patient, but there are important considerations. According to current NCCN guidelines, IRE is generally considered for patients with locally advanced, unresectable pancreatic cancer, especially in cases where the tumor involves major vascular structures, such as the superior mesenteric artery (SMA), as in this case. IRE can be used as a local ablative therapy in patients with no evidence of distant metastasis who are not surgical candidates due to vascular involvement.  However, several key factors must be considered: 1. **Patient Selection**: IRE is most appropriate for patients with locally advanced disease who have completed chemotherapy and/or radiation and have stable or responding disease. 2. **Multimodal Approach**: IRE is often used as part of a multimodal treatment strategy, following systemic chemotherapy to control microscopic metastatic disease. 3. **Tumor Location and Size**: While IRE can be used for tumors encasing vessels, careful evaluation of the tumor's location relative to other vital structures is critical. 4. **Expertise**: IRE requires specialized expertise and should be performed in centers experienced with the technique.  In summary, IRE may be considered for this patient as part of a comprehensive treatment plan after neoadjuvant therapy, but it requires careful multidisciplinary evaluation. | According to the current NCCN guidelines, irreversible electroporation (IRE) is not a standard treatment option for pancreatic ductal adenocarcinoma that encases the superior mesenteric artery. The recommended management for unresectable, locally advanced pancreatic cancer includes systemic chemotherapy, with or without radiation therapy. While IRE is being studied as a potential treatment in clinical trials, its efficacy and safety have not been established to the extent necessary for routine clinical use. Therefore, IRE should only be considered within a clinical trial setting, and standard treatment protocols should be followed. |
| 15 | A 67-year-old patient was diagnosed with resectable pancreatic ductal adenocarcinoma of pancreatic head and scheduled for a pancreaticoduodenectomy. During the procedure, the surgeon used an Endo-GIA stapler to divide the tissue between the superior mesenteric artery and the uncinate process, aiming to reduce operative time and minimize bleeding. Was this approach appropriate? | No, this approach was not appropriate. According to up-to-date guidelines, dividing the tissue between the superior mesenteric artery (SMA) and the uncinate process using a stapler is not recommended. This is a critical area where sharp dissection is usually preferred to ensure negative margins and adequate oncological clearance, especially given the close proximity of vital structures like the SMA. Using a stapler in this region could compromise the oncological integrity of the resection and may lead to inadequate margin clearance. | No, this approach was not appropriate. The area between the superior mesenteric artery and the uncinate process requires meticulous dissection to ensure negative margins and avoid vascular injury. Using an Endo-GIA stapler in this region may compromise oncologic outcomes by increasing the risk of positive margins, which is not aligned with NCCN guidelines. |
| 16 | A 72-year-old patient underwent a pancreaticoduodenectomy for pancreatic head cancer. During the procedure, the surgeon identified enlarged lymph nodes located between the vena cava and the abdominal aorta. Could performing a para-aortic lymph node dissection have potentially enhanced the patient's long-term prognosis? | Performing a para-aortic lymph node dissection in the context of pancreatic cancer is generally not recommended for improving long-term prognosis. According to current NCCN guidelines, para-aortic lymph node metastasis is considered a marker of advanced disease and poor prognosis. The presence of metastases in these nodes typically indicates systemic spread, and dissection does not provide a survival benefit. Therefore, routine para-aortic lymph node dissection is not recommended as it is unlikely to enhance long-term outcomes in this scenario. | Performing a para-aortic lymph node dissection would not have enhanced the patient's long-term prognosis. According to the NCCN guidelines, extended lymphadenectomy—including dissection of para-aortic lymph nodes—has not shown a survival benefit in pancreatic cancer patients undergoing pancreaticoduodenectomy. |
| 17 | A 70-year-old man was diagnosed with pancreatic head cancer following the onset of obstructive jaundice, and a pancreaticoduodenectomy was initially planned. However, during surgery, a nodule was discovered in the greater omentum, and frozen section pathology indicated metastatic adenocarcinoma. As a result, the surgical plan was revised to a choledochojejunostomy. The patient had no contraindications to gastrojejunostomy. Given this, should a gastrojejunostomy be performed concurrently? | Yes, a gastrojejunostomy should be performed concurrently. According to the NCCN guidelines, in patients with pancreatic cancer who are found to have metastatic disease during surgery, palliative bypass procedures such as choledochojejunostomy and gastrojejunostomy are recommended to prevent future gastric outlet obstruction, especially if the patient is at risk. Since the patient has no contraindications, performing a prophylactic gastrojejunostomy is appropriate. | Yes, a gastrojejunostomy should be performed concurrently to prevent future gastric outlet obstruction, as recommended by the NCCN guidelines. |
| 18 | A 75-year-old woman was admitted to the hospital with a diagnosis of pancreatic head cancer. MRI revealed a solitary 2 cm liver lesion, and a pathological biopsy confirmed metastatic pancreatic ductal adenocarcinoma. Could stereotactic body radiation therapy be considered to treat the liver metastasis, followed by a pancreaticoduodenectomy to remove the primary tumor once radiotherapy is completed? | No, stereotactic body radiation therapy (SBRT) for the liver metastasis followed by pancreaticoduodenectomy is not recommended in this case. According to the NCCN guidelines, the presence of metastatic disease, such as a liver lesion confirmed as metastatic pancreatic ductal adenocarcinoma, typically excludes patients from curative surgery like pancreaticoduodenectomy. The primary treatment in metastatic pancreatic cancer would focus on systemic therapy, such as chemotherapy, rather than local treatments like SBRT or surgery. | In patients with pancreatic cancer and confirmed liver metastasis, surgical removal of the primary tumor is generally not recommended. The standard treatment is systemic chemotherapy. Stereotactic body radiation therapy for liver metastasis followed by pancreaticoduodenectomy is not typically advised. |
| 19 | A 69-year-old man was admitted to the hospital with obstructive jaundice. A CT scan revealed a tumor in the head of the pancreas, along with multiple hepatic tumors. A liver biopsy confirmed metastatic pancreatic ductal adenocarcinoma. The patient’s prognosis indicated a life expectancy of approximately six months. In this case, is a plastic biliary stent the optimal choice for biliary drainage? | No, a plastic biliary stent is not the optimal choice for biliary drainage in this case. According to the NCCN guidelines, a metal biliary stent is preferred for patients with metastatic pancreatic cancer and a life expectancy greater than 3-6 months. Metal stents have a longer patency and lower risk of complications compared to plastic stents, making them the better option for palliative management in this context. | No, a plastic biliary stent is not the optimal choice. Given his expected survival of approximately six months, a self-expandable metal stent is preferred due to its longer patency and reduced need for replacements. |
| 20 | A 40-year-old woman was referred for an ultrasound due to pancreatic duct dilation. An MRI revealed that the main pancreatic duct in the body and tail of the pancreas had a maximum diameter of 8 mm, leading to a diagnosis of main-duct intraductal papillary mucinous neoplasm of pancreas. The doctor recommended close surveillence. After five years of follow-up, a subsequent MRI showed that the dilated pancreatic duct had further increased in diameter to 15 mm. Should surgery now be recommended for this patient? | Yes, surgery should be recommended for this patient. According to the NCCN guidelines, a main-duct intraductal papillary mucinous neoplasm (IPMN) with a main pancreatic duct diameter ≥ 10 mm is an indication for surgical resection due to the significantly increased risk of malignancy. Given that the duct has increased to 15 mm, this patient meets the criteria for surgery. | Yes, surgery should now be recommended. In main-duct intraductal papillary mucinous neoplasms (IPMNs), a main pancreatic duct diameter exceeding 10 mm is associated with a higher risk of malignancy. According to current NCCN guidelines, surgical resection is advised when the duct dilates beyond this threshold. |
